# Pharmacotherapy for Alzheimer’s Disease and Dementias in Long-Term Care: A Real-World EHR Study

**DOI:** 10.64898/2026.01.16.25342403

**Authors:** Tyler M. Saumur, Huda Ashraf, Katherine E. Mathers, Brittin L. Wagner

## Abstract

**Objectives:** To characterize contemporary pharmacologic treatment patterns for Alzheimer’s disease and related dementias (ADRD) among U.S. long-term care residents and to examine facility- and resident-level factors associated with treatment.

**Design:** Retrospective, observational study.

**Setting and Participants:** Electronic health record data from 1,675,873 long-term care residents in the PointClickCare Life Sciences database included 359,801 with a documented ADRD diagnosis in skilled nursing facilities in the U.S. (January–April 2025).

**Methods:** Residents were classified as treated/untreated based on receipt of guideline-directed ADRD therapy, consistent with Alzheimer’s Association guidelines. Analyses incorporated demographics, comorbidities, medication burden, and facility characteristics. Multivariate logistic regression estimated odds of receiving guideline-concordant therapy.

**Results:** Overall, 72.5% of residents with ADRD received ≥1 pharmacologic treatment recommended for ADRD. Treatment was most common among residents with Lewy body dementia (83.9%) and early-onset Alzheimer’s disease (82.3%) and least frequent among residents aged ≥90 years (65.1%), Black/African American residents (66.8%), and those with cerebral degeneration (66.8%). Treated residents exhibited higher medication burden (mean 4.4 vs 3.3). Diagnoses for other chronic conditions as well as specific ADRD subtypes strongly impacted probability of treatment; diabetes and hyperlipidemia were associated with lower odds of treatment, whereas ADRD subtypes strongly predicted treatment.

**Conclusions and Implications:** More than one-quarter of residents with ADRD remain untreated with guideline-recommended pharmacotherapy, and treatment varied significantly by non-clinical predictors. These findings underscore the need to investigate and understand possible treatment disparities, optimize polypharmacy management, and discover new ADRD treatments, as current options are often ineffective with many side effects.

**Brief Summary:** This study used real-world data from electronic health records (EHR) to understand treatment patterns of those with Alzheimer’s disease and related dementias (ADRD) in U.S. long-term care facilities. International Classification of Diseases Tenth Revision, Clinical Modification (ICD-10) codes were used to identify ADRD diagnoses and medication orders were used to identify treatment. From January to April 2025, there were 359,801 with a documented ADRD diagnosis in skilled nursing facilities. Over 25% of those with ADRD did not have a medication order for a guideline-recommended pharmacological treatment. Comorbidities of diabetes and hyperlipidemia were associated with lower odds of receiving ADRD treatment, suggesting concerns related to adverse drug reactions and competing clinical priorities. The use of cognitive and disease-modifying therapies was low compared to behavioral/psychiatric medications; this finding suggests a need for more effective and safe drugs that target the root causes of ADRD opposed to the behavioral and psychiatric complications. Taken together, the results of this study call for targeted interventions to address disparities in treatment, enhanced clinical decision-making support regarding polypharmacy, and improved pharmacological options for those with ADRD.

## Introduction

Alzheimer’s disease and related dementias (ADRD) currently affect an estimated 4 million individuals in the United States (U.S.), with numbers projected to quadruple by 2050; ADRD is among the primary causes of dependency and institutionalization in older adults.^1^ Treatment for ADRD can typically be categorized into those that manage behavioral and psychological symptoms, those that manage cognitive symptoms, and those that target disease progression.^2^ While those targeting disease progression such as lecanemab and donanemab are more novel – being approved for early symptomatic Alzheimer’s disease within the last 2 years – drugs treating cognitive symptoms, such as acetylcholinesterase inhibitors (AChEIs) like donepezil and glutamate regulators like memantine, have been approved for more than 20 years. Understanding how these approved treatments for ADRD are used in the long-term care (LTC) setting is important to understanding the context and potential needs to improve treatment.

Since their discovery, pharmacologic treatment using AChEIs and memantine remains inconsistent, particularly in LTC settings. Early data from U.S. LTC facilities demonstrated stark underuse of these medications; in the late 1990s, only about 30% of newly admitted and 19% of long-stay residents with Alzheimer’s disease received donepezil.^3^ A cross-sectional analysis using the 2004 National Nursing Home Survey found that 30% of LTC residents with dementia were being treated with AChEIs, with donepezil representing ~71% of those prescriptions.^4^ In more recent real-world analyses, among 350,197 newly admitted residents with dementia, AChEI and memantine use declined from 44.5% to 36.9% and 27.4% to 23.2% during 2011–2018, respectively.^5^

Among newly admitted LTC residents in the early 2000s, approximately 40% received AChEIs or memantine at the time of admission. In addition, treatment was significantly more common in residents with mild to moderate dementia classified as having Cognitive Performance Scale (CPS) scores of 0 to 4 than in those with advanced/severe disease defined as a CPS score of 5 or 6 (41.2% vs 33.3% of residents, respectively).^6^ By 3 months post-admission, use of AChEIs and memantine was discontinued by 48.6% of residents with mild to moderate dementia and 57% with advanced/severe dementia.^6^ The limited clinically meaningful differences and gastrointestinal complications associated with AChEIs may contribute to their high discontinuation rate.^7,8^ In addition, non-pharmacological interventions such as cognitive stimulation programs may provide a safer alternative that can lead to improvements in communication and social interaction along with modest improvements in cognition.^9^

Treatment gaps reported during the early 2000s are even more striking in assisted living settings (ALFs). In ALFs, cognitive impairment is prevalent in over half of residents (assessed using Mini-Mental State Examination); among those, 63% had no documented dementia diagnosis, and 75% received no AChEIs or memantine.^10^ Community-based data echo these gaps – in one U.S. cohort of mild-to-moderate AD patients, 64.5% had never been treated with an AChEI at the time of evaluation.^1^

Historically documented variations in treatment such as these raise critical questions: what are contemporary treatment rates, and how do they vary by specific ADRD diagnoses and treatment? Treatment inequities documented in past are linked to older age, advanced dementia, under recognition, facility characteristics, and demographic factors.^4,5^ Untreated dementia may accelerate functional decline, increase hospitalizations, and diminish survival.^11^ Accordingly, this study aimed to evaluate up-to-date patterns of pharmacologic treatment for all types of ADRD, all treatments included in ADRD prescribing guidelines, and several facility-level factors in a large electronic health record (EHR) dataset representative of U.S. older adults in LTC.

## Methods

### Setting and Participants

This retrospective study used EHR data from the PointClickCare Life Sciences database, which includes clinical information from over 18 million residents in U.S. LTC facilities including ALFs, senior living, and skilled nursing facilities (SNFs). Exemption from ethics committee oversight was granted by a Research Ethics Board.

Between January and April 2025, there was data available for 1,675,873 LTC residents including 1,251,460 SNF residents. Of these, 359,801 residents had a documented diagnosis of ADRD. Residents were included if they had at least one active ADRD diagnosis, identified by International Classification of Diseases Tenth Revision, Clinical Modification (ICD-10) codes G30.0, G30.1, G30.9, I67.2, F01*, F03*, or G31.83 and at least one day of documented stay in a SNF during the study period. The dataset captured demographics, diagnoses, medication records, and facility characteristics.

### Exposures and Variables of Interest

Residents were categorized as treated or not treated based on the presence of a medication order for any guideline-directed ADRD pharmacologic therapy consistent with the Alzheimer’s Association guidelines.^2^ Treatments included AChEIs (donepezil, rivastigmine, galantamine) and memantine along with other guideline-concordant medications.

Variables of interest included demographic characteristics (age, sex, race/ethnicity), admitting payer type, facility region, and length of stay in the SNF. Days in facility (DIF) was calculated as the number of days between a resident’s admission and discharge dates. For residents in facility at the end of the study period, 30 April 2025 was used as the discharge date. Consecutive visits were combined when separated by gaps of seven days or fewer, treating them as a single continuous stay for DIF calculation. Clinical variables captured comorbid conditions such as diabetes and hyperlipidemia (via ICD-10 code), which have been identified to be linked to medication prescribing patterns in ADRD.

### Data Analysis

Data were extracted using SQL and analyzed in Python. Descriptive statistics were used to summarize resident demographic and clinical characteristics, with means and SDs reported for continuous variables and counts (%). A multivariable logistic regression model estimated the likelihood of receiving guideline-directed ADRD medication, adjusting for demographic, clinical, and facility factors (p <0.001).

## Results

### Demographic and Clinical Characteristics

Overall, there were 359,801 residents with ADRD who met study eligibility criteria, with 260,886 (72.5%) receiving guideline-concordant ADRD medication (Table 1). The mean (SD) age of residents was 80.2 (9.7) years. The majority of residents were female (63.7%), White (69.1%), and from Southern (39.1%) or Midwestern United States (26.8%).

**Table 1.**
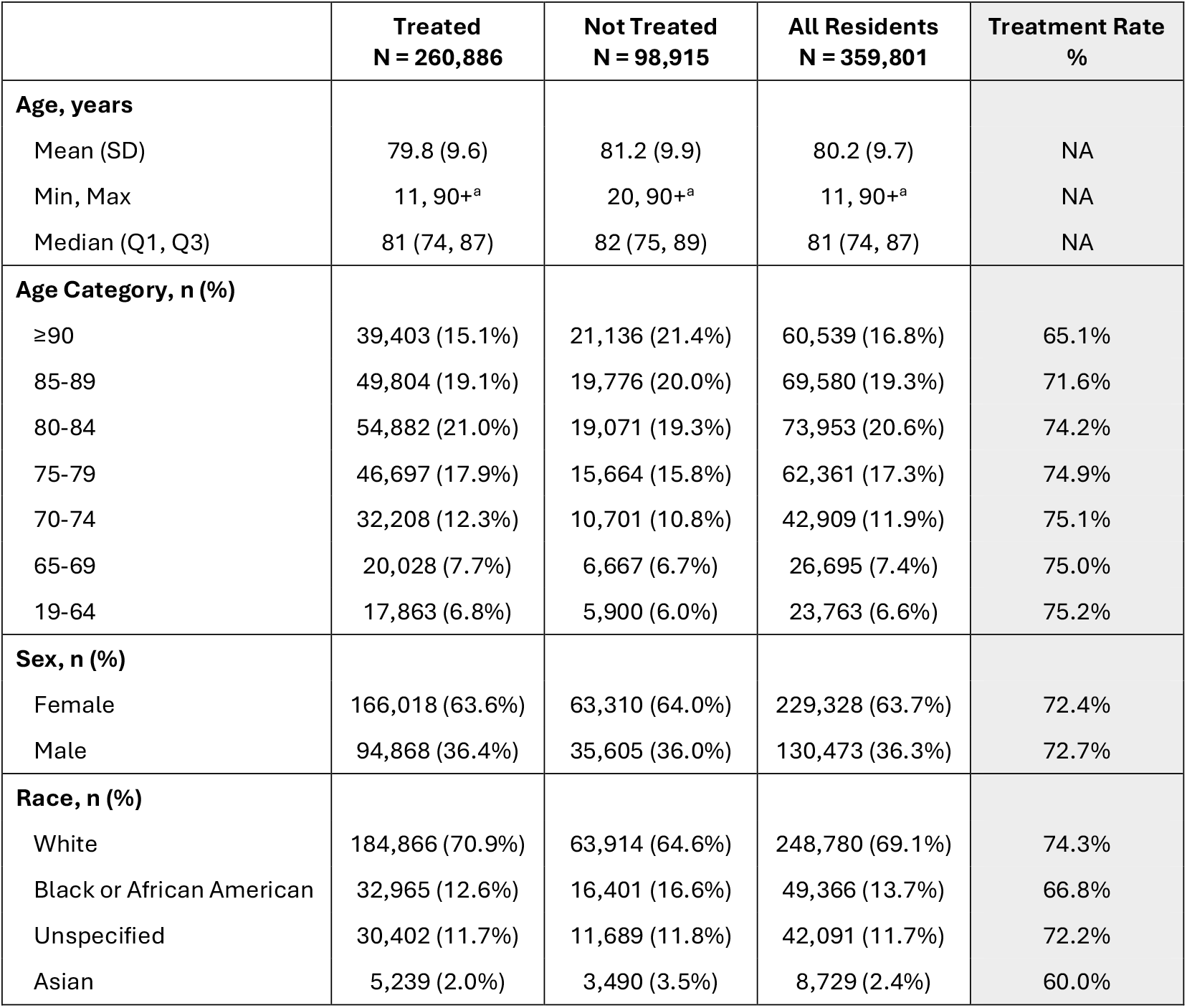
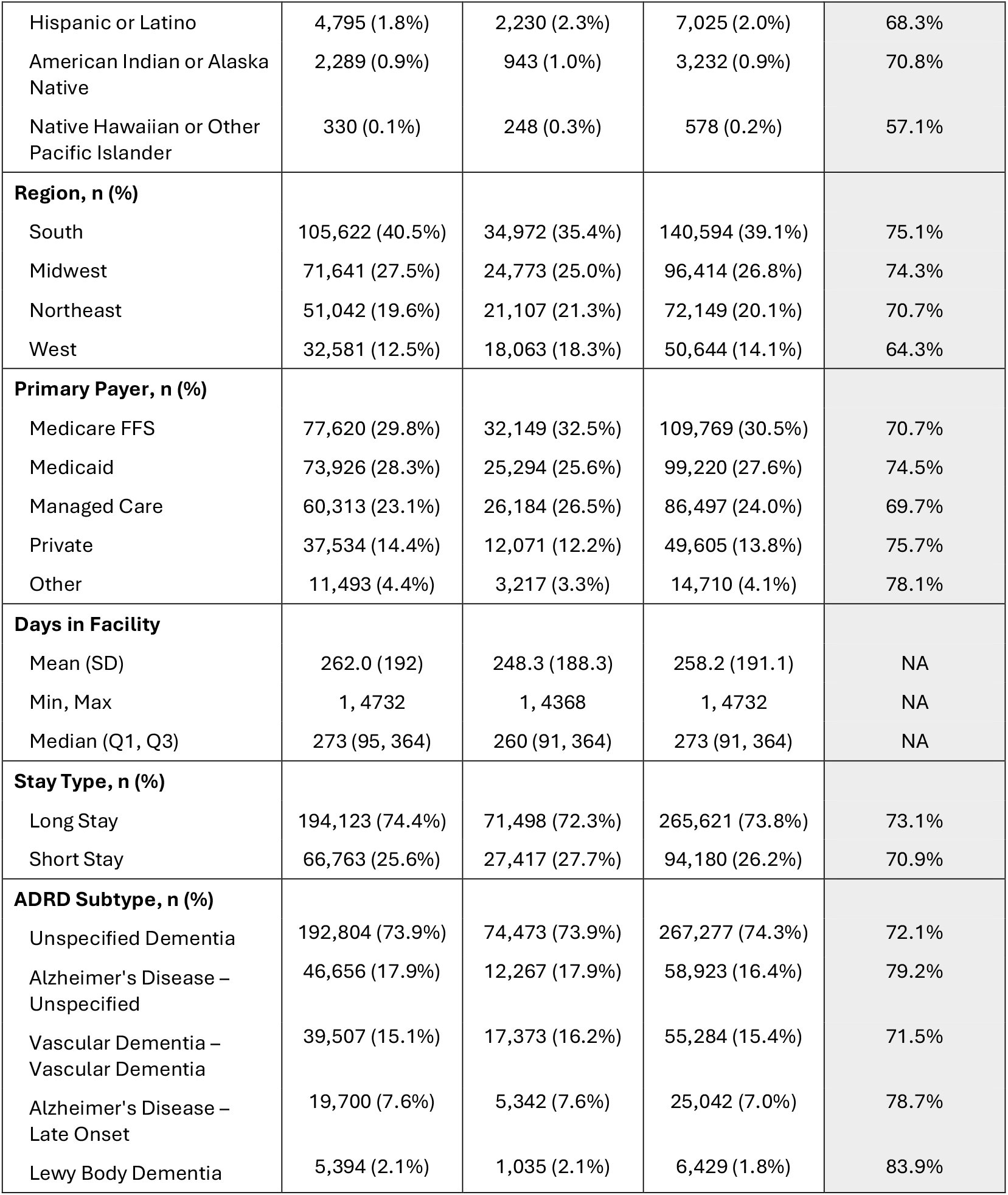
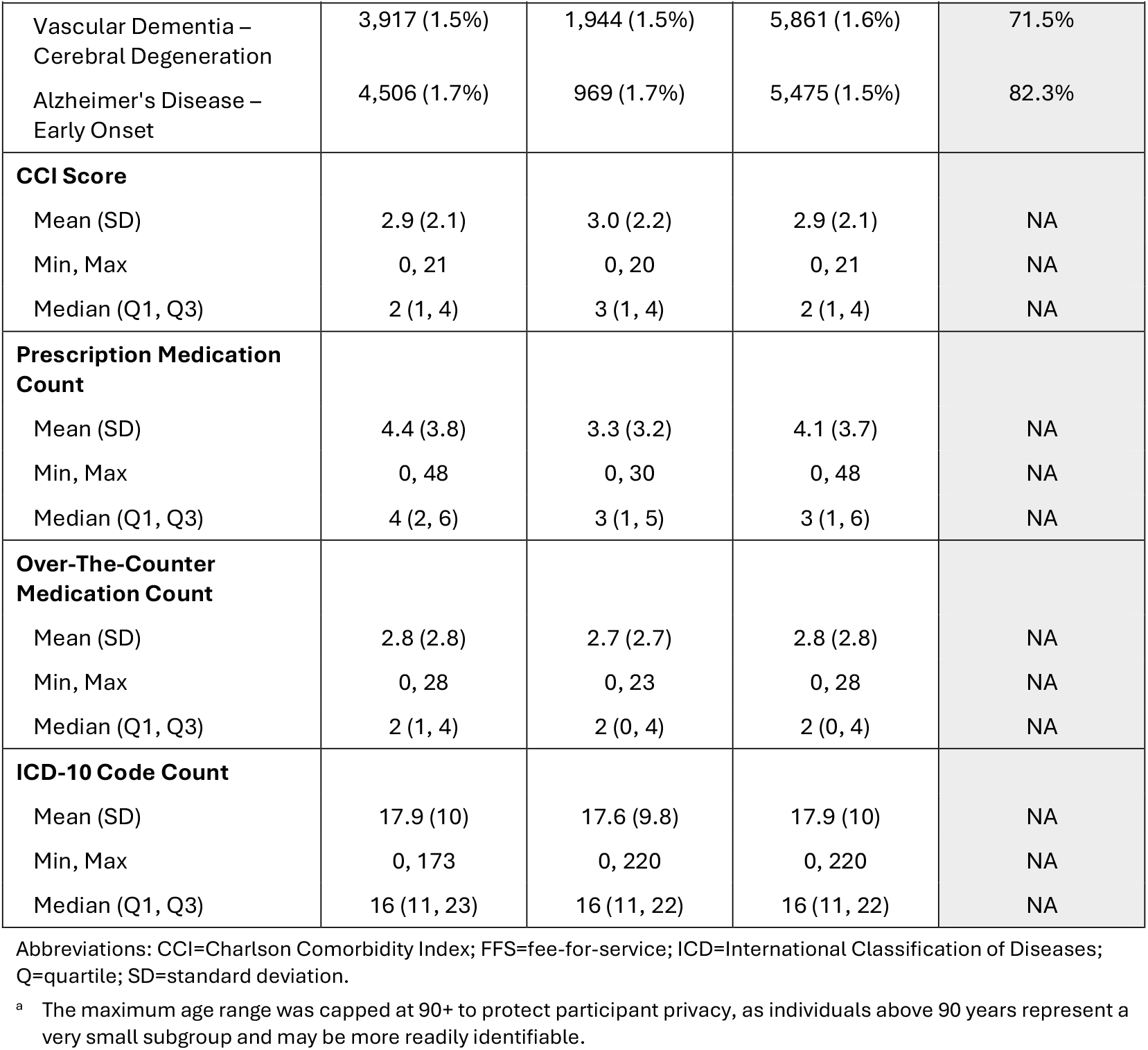
Demographic and Clinical Characteristics Across Exposure Groups.

The most common ADRD diagnoses were unspecified dementia (74.3%), unspecified Alzheimer’s disease (16.4%), and vascular dementia (15.4%; Table 1). The mean (SD) number of ICD-10 codes associated with a resident was 17.9 (10). The mean (SD) CCI score was 2.9 (2.1). The mean (SD) days in facility was 258.2 (191.1) days, ranging from 1 day to ~13 years. The mean (SD) number of prescription and over-the-counterb medications being taken by each resident were 4.1 (3.7) and 2.8 (2.8), respectively. Among all residents with ADRD, the most taken medication classes for ADRD treatment were selective serotonin reuptake inhibitor (SSRIs; 29.6%), AChEIs (25.4%), atypical antipsychotics (22.6%), and benzodiazepines (21.2%; Table 2).

**Table 2.** Treatment Rates by Medication Class.

|  | **Treated**<br>**N = 359,801** |
| --- | --- |
| **Medication Class, n (%)** |  |
| Selective Serotonin Reuptake Inhibitors | 106,518 (29.6%) |
| Acetylcholinesterase Inhibitors | 91,350 (25.4%) |
| Atypical Antipsychotics | 81,480 (22.6%) |
| Benzodiazepines | 76,161 (21.2%) |
| NMDA Receptor Antagonists | 66,845 (18.6%) |
| Serotonin Antagonist and Reuptake Inhibitors | 55,338 (15.4%) |
| Anticonvulsants | 6,956 (1.9%) |
| Sedative-Hypnotics | 2,172 (0.6%) |
| Novel Disease-Modifying Therapies (Anti-Amyloid Monoclonal Antibodies) | 1 (0.0%) |

Abbreviations: NMDA=N-methyl-D-aspartate.

### Treatment Rates and Odds Ratios

Treatment rates were highest for residents with Lewy body dementia (83.9%), early onset Alzheimer’s disease (82.3%), unspecified Alzheimer’s disease (79.2%), late onset Alzheimer’s disease (78.7%), and those with an admitting payer type of “Other” (78.1%; Table 1). Treatment rates were the lowest for residents who were Native Hawaiian or Other Pacific Islander (57.1%), Asian (60.0%), from Western United States (64.3%), ≥90 years of age (65.1%), Black or African American (66.8%), and had cerebral degeneration (66.8%). The mean (SD) number of total active prescription medications was 4.4 (3.8) for those treated compared to 3.3 (3.2) for those not treated. Other baseline characteristics were comparable between the two groups.

Odds ratios were generated following a multivariate logistic regression model, which comprised of 67 variables (Supplementary Table 1). Odds ratios ranged from 0.74 to 2.89, with the 5 highest and lowest odds ratios displayed in Figure 1. Unsurprisingly, the top variables with statistically significant odds ratios associated with receiving ADRD guideline-concordant medication were the specific ADRD subtype diagnoses. Variables with the lowest statistically significant odds ratios were typically related to diabetes and hyperlipidemia diagnoses and medications.

**Figure 1.**
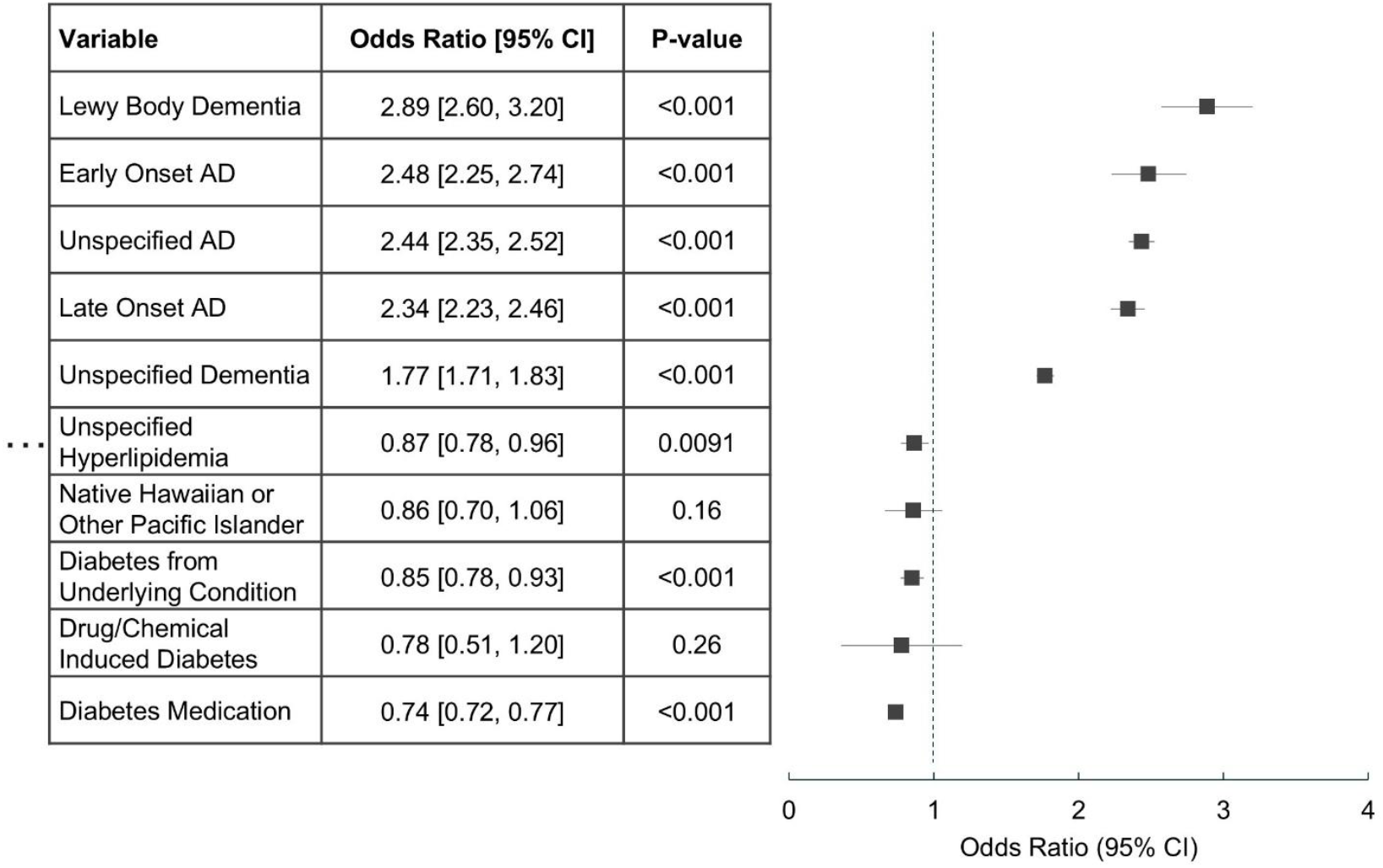
Five Highest and Lowest Odds Ratios. Footnote: Abbreviations: AD=Alzheimer’s disease; CI=confidence interval

## Discussion

This study found that of 359,801 SNF residents with ADRD in the U.S., 27.5% are not receiving guideline-concordant pharmacologic treatment, highlighting a substantial gap in Alzheimer’s disease and dementia care. Treatment rates varied considerably by ADRD diagnosis subtype, with residents diagnosed with Lewy body dementia having the highest likelihood of receiving therapy. This may reflect greater clinical recognition of neuropsychiatric symptoms as well as treatment responsiveness of hallmark symptoms in this subgroup, as suggested in prior studies.^12,13^

Lower treatment rates were observed among residents also diagnosed with diabetes and/or hyperlipidemia. This finding may be due to concerns around adverse drug interactions, increased risk of side effects, competing clinical priorities in the context of multiple chronic conditions, and myriad other potential drivers, generating powerful hypotheses to be tested empirically. Treated residents had a significantly higher number of prescription medications compared to untreated residents (mean prescription medication count of 4.4 vs 3.3), raising important considerations regarding medication burden, prescriber behavior and resident preferences, and the potential for drug-drug interactions. As polypharmacy is known to have an independent negative impact on cognition, treated individuals with ADRD may also be at a further risk of cognitive impairment as a result of polypharmacy.^14^

The treatment rate for first-line medications such as AChEIs was only 25.4%, which aligns with treatment rates observed in the community and LTC setting in the U.S. as well as LTC facilities in Sweden.^15–17^ Interestingly, the treatment rates from the U.S. were reported over 20 years ago, suggest little change in front-line ADRD treatment rates over time.^15,16^ These treatment rates likely reflect clinical decision-making surrounding the costs and benefits of prescribing AChEIs, which still require higher quality evidence regarding their ability to reduce disease severity and maintain cognition, along with formulations with better safety profiles.^7,8,18^ Furthermore, the most common types of treatment used for ADRD were behavioral/psychiatric management drugs such as SSRIs, antipsychotics, and benzodiazepines, with <1% of residents receiving novel disease-modifying therapies. These findings further highlight the need for more research into disease-modifying therapies that can target the pathophysiology of the disease with less reliance on managing behavioral/psychiatric symptoms.

Limitations of this study include lack of information on clinical appropriateness, medications and treatments historically used by residents that may have informed current choices, and resident preferences more generally. Another limitation is the missing or incomplete data prior to admission to the LTC setting; ADRD diagnoses were dependent on the ICD-10 codes documented in the EHR database, which may often be generalized as observed in the present study with 74.3% of residents with ICD-10 codes for “unspecified dementia”. Lastly, the study’s findings may be most applicable to similar LTC settings in the U.S. but may not be as translatable to other healthcare settings, health systems, and countries with different patient demographics and care practices. Further details including historical medication usage among residents, and LTC resident discharge information could help inform generalizability (for example, among residents discharged to community).

## Conclusions and Implications

The findings of this study underscore significant, ongoing gaps in the delivery of guideline-concordant pharmacologic treatment for SNF residents with ADRD, with over a quarter of this population not receiving recommended therapies. These results highlight the need for targeted interventions to address disparities in treatment, particularly among residents with comorbidities such as diabetes and hyperlipidemia. The observed association between polypharmacy and increased treatment rates also raises important considerations for balancing therapeutic benefit with the risks of medication burden and drug interactions. From a practice and policy perspective, these insights call for enhanced clinical decision-making support, increased provider education, and system-level strategies to ensure equitable and appropriate dementia care. Future research should focus on developing and evaluating interventions to target ADRD pathophysiology with a favorable safety profile, optimize medication management, reduce disparities, and carefully weigh the benefits and risks of polypharmacy in this vulnerable population.

## Data Availability

Data produced in the present study are available upon reasonable request to the authors.

## Acknowledgements

We would like to thank Jody Long and Dr. Steve Buslovich for their collaboration and their clinical input provided for this manuscript. This research was supported by PointClickCare Life Sciences and McMaster University. McMaster University Library provided journal article access.

---

**Supplementary Table 1.** Complete List of Variables and Odds Ratios in the Multivariate Logistic Regression Model.

| Variable (ICD-10 Codes) | Odds Ratio [95% CI] | P-value |
| --- | --- | --- |
| Lewy Body Dementia (G31.83) | 2.89 [2.60, 3.20] | <0.001 |
| Alzheimer's Disease - early onset (G30.0) | 2.48 [2.25, 2.74] | <0.001 |
| Alzheimer's Disease - unspecified (G30.9) | 2.44 [2.35, 2.52] | <0.001 |
| Alzheimer's Disease - late onset (G30.1) | 2.34 [2.23, 2.46] | <0.001 |
| Unspecified Dementia (F03*) | 1.77 [1.71, 1.83] | <0.001 |
| Depression diagnosis (any) active during current visit | 1.67 [1.53, 1.82] | <0.001 |
| Vascular Dementia - vascular dementia (F01*) | 1.48 [1.43, 1.53] | <0.001 |
| Depression - Unspecified (F32.A) | 1.48 [1.36, 1.60] | <0.001 |
| Depression - MDD (F33, F33.0, F33.1, F33.2, F33.3, F33.4, F33.40, F33.41, F33.42) | 1.42 [1.31, 1.54] | <0.001 |
| CMS ownership type (converted to binary from categorical) | 1.36 [1.31, 1.42] | <0.001 |
| Depression - Other (F33.8) | 1.35 [1.22, 1.50] | <0.001 |
| Hispanic or Latino | 1.34 [1.25, 1.43] | <0.001 |
| White | 1.30 [1.26, 1.34] | <0.001 |
| Midwest | 1.28 [1.24, 1.33] | <0.001 |
| Count of Rx med orders active at Index Date | 1.22 [1.22, 1.23] | <0.001 |
| Vascular Dementia - cerebral degeneration (I67.2) | 1.20 [1.11, 1.30] | <0.001 |
| Southern United States | 1.16 [1.12, 1.20] | <0.001 |
| CMS ownership type (converted to binary from categorical) | 1.15 [1.10, 1.20] | <0.001 |
| Payer – Private | 1.14 [1.08, 1.20] | <0.001 |
| American Indian or Alaska Native | 1.13 [1.01, 1.25] | 0.027 |
| Northeastern United States | 1.12 [1.08, 1.16] | <0.001 |
| SVI - Socioeconomic percentile (for facility county) | 1.08 [0.99, 1.19] | 0.097 |
| Payer – Medicaid | 1.07 [1.02, 1.13] | 0.007 |
| Resident sex (converted to binary from categorical) | 1.07 [1.05, 1.10] | <0.001 |
| Hyperlipidemia diagnosis (any) active during current visit | 1.06 [0.96, 1.18] | 0.26 |
| Medication order for any Hyperlipidemia med during current visit | 1.05 [1.03, 1.08] | <0.001 |
| SVI - Household characteristics percentile (for facility county) | 1.03 [0.98, 1.08] | 0.27 |
| Resident average ADL scores (daily ADLs) | 1.02 [1.02, 1.02] | <0.001 |
| Facility Staffing Rating (from CMS Provider Info) | 1.02 [1.01, 1.03] | <0.001 |
| Resident current BMI | 1.01 [1.01, 1.01] | <0.001 |

| Variable (ICD-10 Codes) | Odds Ratio [95% CI] | P-value |
| --- | --- | --- |
| Days since Dx onset | 1.00 [1.00, 1.00] | 0.59 |
| Facility Avg number of residents per day (from CMS Provider Info) | 1.00 [1.00, 1.00] | <0.001 |
| Facility health inspection rating (from CMS Provider Info) | 1.00 [0.98, 1.02] | 0.96 |
| Facility number of CMS-certified beds (from CMS Provider Info) | 1.00 [1.00, 1.00] | <0.001 |
| Facility Quality Measures rating (from CMS Provider Info) | 1.00 [0.99, 1.01] | 0.92 |
| Days since Dx onset | 1.00 [1.00, 1.00] | <0.001 |
| Days since Dx onset | 1.00 [1.00, 1.00] | 0.02 |
| Type 1 diabetes mellitus (E10%) | 1.00 [0.87, 1.15] | 0.96 |
| Days in facility | 1.00 [1.00, 1.00] | <0.001 |
| Days since Dx onset | 1.00 [1.00, 1.00] | 0.003 |
| Days since Dx onset | 1.00 [1.00, 1.00] | <0.001 |
| Facility bed count | 1.00 [1.00, 1.00] | <0.001 |
| SVI - Racial/minority percentile (for facility county) | 1.00 [1.00, 1.00] | 0.20 |
| Resident age at index date | 0.99 [0.99, 0.99] | <0.001 |
| Count of diagnoses (unique ICD-10 codes) active at Index Date | 0.99 [0.99, 0.99] | <0.001 |
| Rural Urban Code | 0.99 [0.99, 1.00] | <0.001 |
| Count of OTC med orders active at Index Date | 0.98 [0.98, 0.98] | <0.001 |
| Facility Five-star rating (from CMS Provider Info) | 0.97 [0.95, 0.99] | 0.004 |
| Type 2 diabetes mellitus E11% | 0.97 [0.86, 1.10] | 0.62 |
| SVI - Housing/transportation percentile (for facility county) | 0.97 [0.93, 1.02] | 0.30 |
| Hypertension diagnosis (any) active during current visit | 0.96 [0.93, 0.99] | 0.005 |
| Average BIMS score | 0.95 [0.95, 0.95] | <0.001 |
| Medication order for any Hypertension med during current visit | 0.95 [0.93, 0.98] | <0.001 |
| Payer – Medicare FFS | 0.95 [0.90, 1.00] | 0.05 |
| Asian | 0.93 [0.88, 0.99] | 0.03 |
| Diabetes diagnosis (any) active during current visit | 0.92 [0.81, 1.05] | 0.184 |
| SVI - Overall percentile (for facility county) | 0.92 [0.84, 1.01] | 0.10 |
| Charleston Comorbidity Index Score | 0.91 [0.91, 0.92] | <0.001 |
| Payer - Managed Care | 0.91 [0.87, 0.96] | <0.001 |
| Black or African American | 0.91 [0.88, 0.95] | <0.001 |
| Hyperlipidemia - mixed (E78.2) | 0.90 [0.82, 0.98] | 0.003 |

| Variable (ICD-10 Codes) | Odds Ratio [95% CI] | P-value |
| --- | --- | --- |
| Other specified diabetes mellitus (E13%) | 0.88 [0.78, 0.99] | 0.003 |
| Hyperlipidemia - Unspecified (E78.5) | 0.87 [0.78, 0.96] | 0.0091 |
| Native Hawaiian or Other Pacific Islander | 0.86 [0.70, 1.06] | 0.16 |
| Diabetes mellitus due to underlying condition (E08%) | 0.85 [0.78, 0.93] | <0.001 |
| Drug or chemical induced diabetes mellitus (E09%) | 0.78 [0.51, 1.20] | 0.26 |
| Medication order for any Diabetes med during current visit | 0.74 [0.72, 0.77] | <0.001 |

